# Machine Learning for Predicting and Maximizing the Response of Breast Cancer Patients to Neoadjuvant Therapy

**DOI:** 10.1101/2025.10.11.25337587

**Authors:** Janie Y. Cai, Zhibin Chen

**Affiliations:** Miami Palmetto Senior High School, Miami, FL; Department of Microbiology and Immunology, Sylvester Comprehensive Cancer Center, University of Miami Miller School of Medicine, Miami, FL

## Abstract

**Purpose:** Neoadjuvant therapy (NAT) is an established treatment for certain high-risk, locally advanced, or unresectable breast cancers, often facilitating breast-conserving surgery. Recent studies show that achieving pathologic complete response (pCR) after NAT correlates with higher event-free survival rates. Thus, accurate prediction of pCR is essential for personalizing breast cancer (BC) treatment to minimize side effects and improve effectiveness.

**Methods:** We used a machine learning model named XGBoost to predict pCR in BC patients. The classifier was trained on the expression values of 4,000 genes to predict pCR in ten arms of the I-SPY2 clinical trial. Based on these predictions, we developed a strategy to maximize pCR likelihood and identified influential genes using the importance scores from the model.

**Results:** XGBoost models for three arms, Pembrolizumab, ABT 888 plus carboplatin, and T-DM1 plus pertuzumab, achieved the highest prediction accuracies with areas under the receiver operation characteristic curve (AUCs) of 0.814, 0.792, and 0.788, respectively. If treatment assignments followed the XGBoost predictions, pCR rates for nine out of ten I-SPY2 arms could increase significantly, by 9.9% to 29.1% compared to trial results. Key genes associated with pCR were identified for each arm. The expression levels of some genes, including lower expression of ABDH1, AMZ1, BAIAP3 and SYTL4 and higher expression of DENND1C, HMGB3, HMMR, PLEKHF1 and RASEF, were associated with pCR in multiple arms.

**Conclusion:** The machine learning models developed in this study provide accurate pCR predictions, improve pCR rates, and may find clinical applications to enhance the treatment for BC patients.

## Introduction

Traditionally, systemic therapy for breast cancer (BC) patients has been administrated following surgery. However, neoadjuvant therapy (NAT), which involves providing systemic treatment prior to surgery, has been found as equally effective for overall survival and long-term recurrence rates when compared to adjuvant therapy (1, 2). NAT can reduce the size of the primary tumor, which leads to surgical resection becoming more manageable, converting inoperable tumors to resectable ones, and aiding in breast-conserving surgery (BCS) (3). Additionally, it offers vital insights into the tumor’s response to treatment, which can guide post-operative decision-making and long-term prognostic assessments (3, 4). NAT has been an established therapeutic approach for certain high-risk, locally advanced or unresectable breast cancers and is often employed to enhance BCS feasibility (5, 6).

NAT also offers a potentially more effective trial design for assessing the efficacy of novel therapies for BC by using pathological complete response (pCR) as an endpoint, in contrast to the conventional drug development process reliant on large phase III trials (7). Randomized neoadjuvant studies indicate that pCR, which is characterized by the absence of disease in both breast tissue and lymph nodes, may be a predictive marker for long-term outcomes in patients with early-stage BC (8). Following this evidence, the US Food and Drug Administration provided guidance endorsing pCR as a primary endpoint to facilitate accelerated drug approval (9). More recent studies have also demonstrated that achieving pCR is associated with significantly better event-free survival (EFS) and overall survival (OS) (6, 10).

The I-SPY2 neoadjuvant platform trial is a clinical study aimed at swiftly identifying new drugs and combinations for treating high-risk and early-stage breast cancer that demonstrate greater efficacy than standard chemotherapy (11). Patients receive the experimental treatment prior to surgery to allow for rapid assessment of tumor response. The trial utilizes the analysis of biomarkers found in a patient’s tumor to determine which treatment is likely to be most effective for them. I-SPY2 provides a comprehensive data set that includes pretreatment gene expression profiles, tumor epithelium-specific protein and phosphoprotein data, as well as clinical response information from BC patients (12). From data involving 987 BC patients across 10 trial arms, five distinct subtypes of breast cancer have been established, forming the RPS-5 response-predictive subtyping schema (12). It was found that if patients were assigned to their optimal treatment based on the RPS-5 classifications, the overall pCR rate would rise to 58% across the nine experimental arms, compared to the original pCR rate of 35% in the same arms and just 19% in the standard-of-care control arm with paclitaxel.

In this study, we employed a machine learning (ML) approach to predict the likelihood of pCR in BC patients undergoing neoadjuvant therapy, utilizing gene expression and clinical data from the I-SPY2 trial (12). Although several studies have used machine learning methods to predict pCR in BC patients following NAT (13-18), gene expression data from the I-SPY2 trial have not been explored in these studies. In our study, we demonstrated that the pCR rate can be significantly enhanced across nine out of ten I-SPY2 arms by leveraging the predictions generated by the ML model. Utilizing these pCR predictions, we then devised a strategy to optimize the number of patients achieving pCR. We also identified specific genes associated with pCR pertinent to each drug combination tested in the experimental arms.

## Methods

### Data set

Microarray gene expression data of 987 BC patients from 10 arms of I-SPY2 were downloaded from the Gene Expression Omnibus (GEO) with the accession number GSE194040. The clinical data of these patients in Table S2 of (12) are downloaded from the journal website that hosts the paper. The total number of patients and the number of patients with pCR in each arm and in the four categories, defined by the hormone receptor (HR) and the human epidermal growth factor receptor 2 (HER2), are extracted from the clinical data and summarized in **Table 1**. The gene expression data set contains the expression values of 19,134 genes.

**Table 1.**
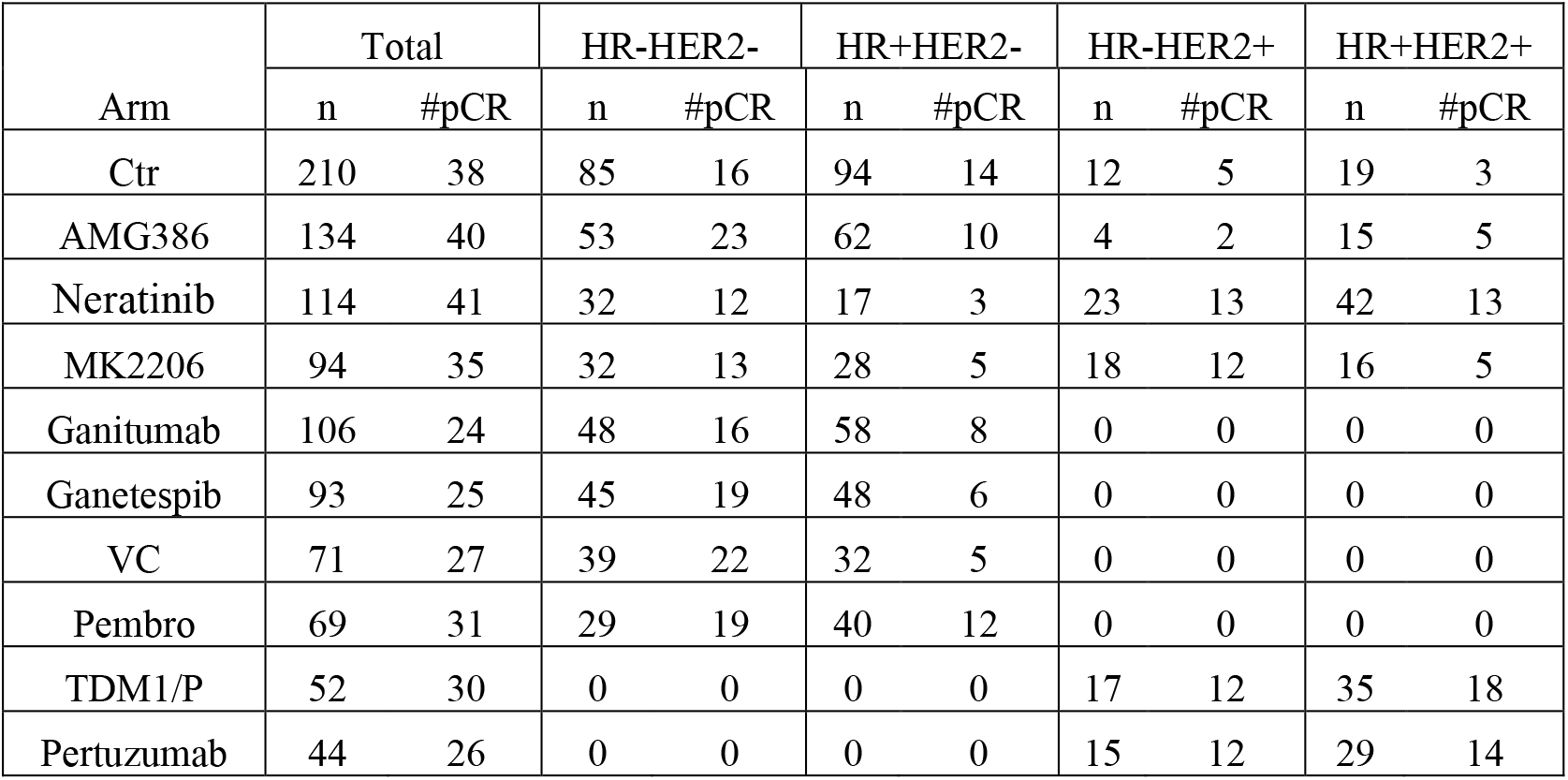
Number of patients in each arm of I-SPY2 (12)

| Arm | Total |  | HR-HER2- |  | HR+HER2- |  | HR-HER2+ |  | HR+HER2+ |  |
| --- | --- | --- | --- | --- | --- | --- | --- | --- | --- | --- |
|  | n | #pCR | n | #pCR | n | #pCR | n | #pCR | n | #pCR |
| Ctrl | 210 | 38 | 85 | 16 | 94 | 14 | 12 | 5 | 19 | 3 |
| AMG386 | 134 | 40 | 53 | 23 | 62 | 10 | 4 | 2 | 15 | 5 |
| Neratinib | 114 | 41 | 32 | 12 | 17 | 3 | 23 | 13 | 42 | 13 |
| MK2206 | 94 | 35 | 32 | 13 | 28 | 5 | 18 | 12 | 16 | 5 |
| Ganitumab | 106 | 24 | 48 | 16 | 58 | 8 | 0 | 0 | 0 | 0 |
| Ganetespib | 93 | 25 | 45 | 19 | 48 | 6 | 0 | 0 | 0 | 0 |
| VC | 71 | 27 | 39 | 22 | 32 | 5 | 0 | 0 | 0 | 0 |
| Pembro | 69 | 31 | 29 | 19 | 40 | 12 | 0 | 0 | 0 | 0 |
| TDM1/P | 52 | 30 | 0 | 0 | 0 | 0 | 17 | 12 | 35 | 18 |
| Pertuzumab | 44 | 26 | 0 | 0 | 0 | 0 | 15 | 12 | 29 | 14 |

After removing genes with missing values, 17,997 genes were left. We then calculated the variance of each gene across 987 samples. The largest variance is 13.72. We ranked the genes according to their variance, and extracted expression values of top 4,000 genes that would be used to train machine learning models for predicting pCR. Of note, the smalles variance of these 4,000 gene is 0.53. The variance of other genes is relatively small, and thus, they may not have much predictive power for pCR and were discarded. The 4,000 genes include HER2 (also known as ERBB2), estrogen receptor 1 and 2 (ESR1 and ESR2), and the progesterone receptor (PGR).

### Machine Learning Method

The statues of HR and HER2 in the clinical data were added to expression values of 4,000 genes as features, which along with the class label of each patient indicating whether the patient achieved pCR were used to train a classifier named XGBoost (19) for each arm of I-SPY2. Specifically, the gene expression and pCR data from an arm was randomly split into a training set with 80% samples and a test set with 20% samples. The training set was used to train an XGBoost classifier. Five-fold cross validation was carried out to determine the optimal values of several hyper-parameters including the number of trees, the maximum depth of a tree, the learning rate, colsample_bynode, and the *l*_2_-regularization weight. The value of scale_pos_weight was set to be the ratio of the count of negative instances to the count of positive instances to mitigate the possible problem of data imbalance between the two classes. Finally, an XGBoost classifier was trained on the whole training data set with the optimal values of hyper-parameters. The test data were then applied to the trained model to evaluate the performance of the model.

The receiver operating characteristic (ROC) curve, the area under the ROC curve (AUC), the precision-recall (PR) curve, and the area under the PR curve (AUPRC) were calculated as the performance metrics. The class label was predicted based on the class probability. The optimal cutoff probability for predicting the class label was determined using Youden’s J statistics (20). Specifically, after a model was trained in CV, the validation set was used to obtain a ROC curve, and the optimal cutoff probability was calculated based on the ROC using the Youden’s J statistic. Since five-fold CV was used, the final optimal cutoff probability was the average of the five cutoff values obtained with five validation folds. After class labels were predicted, accuracy, sensitivity, specificity, precision, and recall were calculated for performance evaluation. The procedure of randomly splitting data, training and testing a classifier was repeated 20 times, and the average performance metrics and their standard deviations were calculated. Of note, the test data were never used in model training.

### Identification and Analysis of Genes Associated with pCR

Each trained XGBoost classifier outputs an importance score for each feature. We ranked genes according to their importance scores and each was assigned a rank from one to 4,000. As described earlier, the process of random data splitting and model training was repeated 20 times. Therefore, each gene had 20 ranks. The outlier ranks of each gene were identified and removed. More specifically, the interquartile range (IQR) of the ranks of each gene was calculated, and any rank that is greater than 1.5*IQR was determined as outliers and discarded. The average rank of each gene was calculated, and finally all genes were re-ranked according to their average ranks. Top genes were deemed to have significant influence on pCR.

We took the top 100 genes for each arm of ISPY-2 and performed gene ontology (GO) analysis. The set of 100 genes was input to GOrilla, an online tool for GO analysis (21), as the target set, and similarly the set of all other human protein-coding genes was input to GOrilla as the background set. Gorilla outputs a directed acyclic graph (DAG) of GO terms related to the 100 genes. Any GO term at any leaf node of the DAG with a p-value less than 10^-3^ was identified as a term significantly enriched in the gene set.

### Maximize pCR

Since we can predict a patient’s pCR to each drug for which an XGBoost model has been trained, each patient can have personalized treatment by choosing the most suitable drug based on pCR predictions, which may maximize the likelihood of pCR. To test this idea, we set aside 324 patients in the two arms Ctr and Neratinib as test samples, then applied the eight XGBoost models trained for the remaining eight arms to these test samples. Thus, each patient in the test samples had eight probabilities of pCR to the drugs in the eight arms. If the probability for a drug is greater than a threshold, then the drug is a candidate treatment for the patient. The probability threshold was determined as follows. As described earlier, after an XGBoost model was trained, we used the test data to compute a PR curve by calculating precisions and recalls for a set of probability thresholds. In this process, we also obtained a curve of precision versus probability threshold, which is named as the PPT curve. Note that precision is the ratio between the true positive and the predicted positive, which is identical to the pCR rate for the drug. Given a target pCR rate, say 0.7, we can determine the probability threshold for each drug based on its PPT curve. After candidate drugs are determined for a patient, a treatment decision can be made by selecting the drug with the highest probability of pCR or the least potential side effect for the patient.

## Results

### ML Models Improve the pCR Rate

We trained an XGBoost classifier for each of 10 arms of the I-SPY2 trial to predict pCR. **Figure 1** illustrates the ROC curves for these classifiers. The XGBoost models for Pembro, VC, and TDM1/P achieved the highest AUCs, with values of 0.81, 0.79, and 0.79, respectively. Conversely, the model for MK2208 attained the lowest AUC at 0.58, while the AUCs of classifiers for other six arms are between 0.62 and 0.71. **Table 2** summarizes the AUC, prediction accuracy, sensitivity, specificity, and F1-score of XGBoost classifiers across all 10 arms of the I-SPY2 trial. Once again, models for Pembro, VC, and TDM1/P achieved the best performance in terms of the F1-score and sensitivity. The model for Ctr demonstrated the lowest F1-score, mainly due to its limited sensitivity. Although it achieved an accuracy of 0.702, which is relatively high, the significant imbalance in sample sizes between the two classes—as shown in Table 1—suggests that accuracy may not adequately reflect the model’s performance compared to the F1-score. PR curves for the ten XGBoost models are presented in S1 Fig. The models for TDM1/P, Pembro, Pertuzumab, and VC achieved highest AUPRCs of 0.69, 0.64, 0.62, and 0.61, respectively.

**Table 2.** Performance of XGBoost classifiers in predicting pCR for 10 I-SPY2 arms.

| Arm | AUC | Accuracy | Sensitivity | Specificity | F1 score |
| --- | --- | --- | --- | --- | --- |
| Ctr | 0.622±0.097 | 0.702±0.078 | 0.419±0.191 | 0.767±0.121 | 0.334±0.104 |
| AMG386 | 0.713±0.077 | 0.683±0.069 | 0.569±0.183 | 0.732±0.118 | 0.505±0.115 |
| Neratinib | 0.653±0.071 | 0.632±0.063 | 0.565±0.198 | 0.677±0.149 | 0.537±0.109 |
| MK2206 | 0.578±0.122 | 0.595±0.091 | 0.481±0.182 | 0.665±0.138 | 0.464±0.139 |
| Ganitumab | 0.670±0.090 | 0.648±0.081 | 0.590±0.214 | 0.665±0.139 | 0.423±0.099 |
| Ganetespib | 0.711±0.122 | 0.705±0.092 | 0.510±0.257 | 0.775±0.149 | 0.449±0.177 |
| VC | 0.792±0.086 | 0.713±0.079 | 0.708±0.189 | 0.717±0.143 | 0.656±0.107 |
| Pembro | 0.814±0.094 | 0.768±0.090 | 0.725±0.199 | 0.800±0.115 | 0.718±0.133 |
| TDM1/P | 0.788±0.132 | 0.691±0.123 | 0.707±0.178 | 0.662±0.163 | 0.734±0.133 |
| Pertuzumab | 0.671±0.179 | 0.594±0.154 | 0.610±0.300 | 0.575±0.238 | 0.590±0.222 |

**Figure 1.**
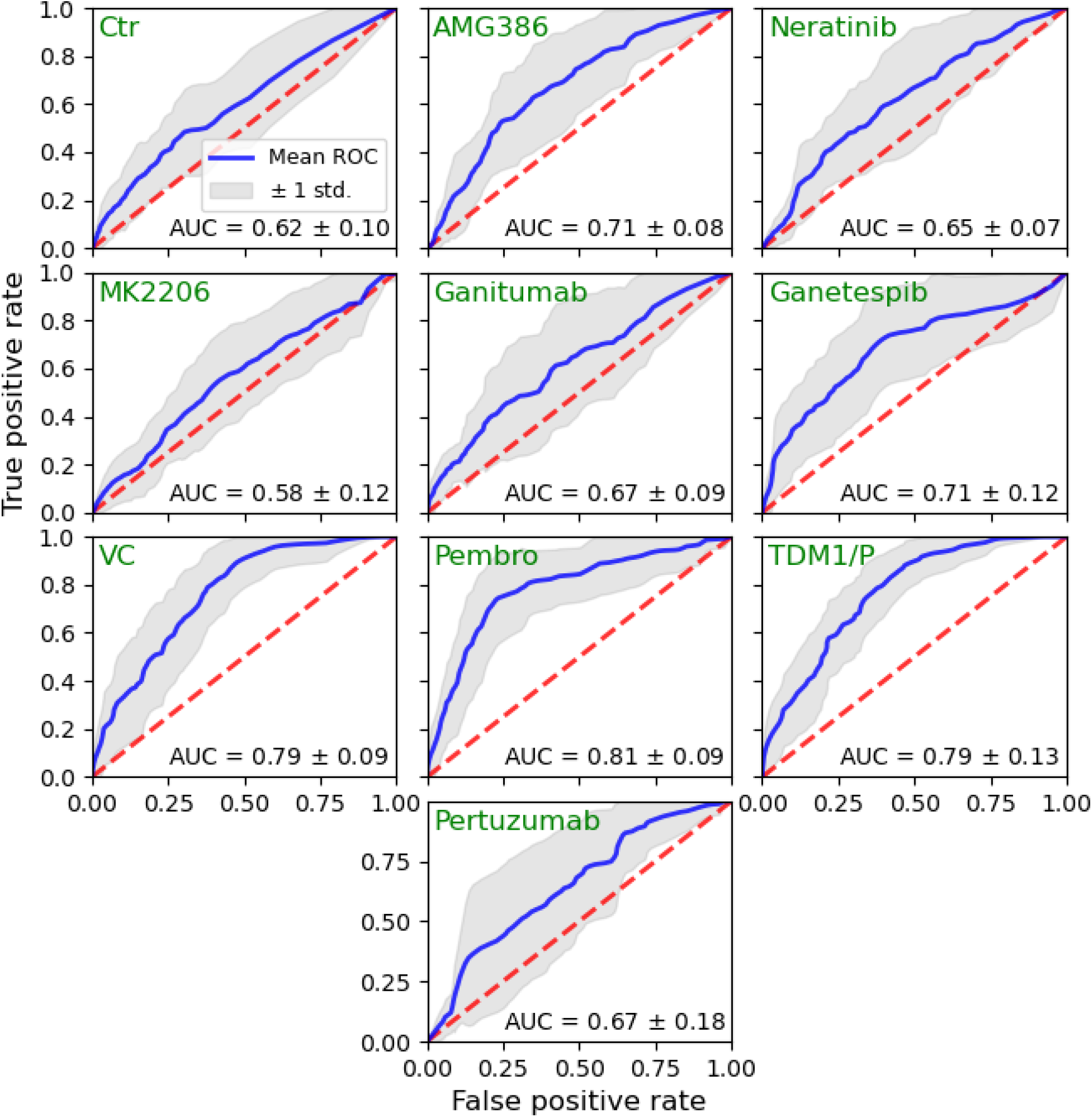
ROC curves of XGBoost classifiers for predicting the pCR in ten arms of I-SPY2.

**Table 3** compares the pCR rate of each I-SPY2 arm, calculated from the data in Table 1, with the precision of each XGBoost model, which is equivalent to the pCR rate of the arm if patients were predicted by the XGBoost gene expression model to have pCR to a treatment regimen and then treated with that regimen. Using the prediction from the XGBoost models, all arms, except for Pertuzumab, exhibit significantly higher pCR rates than those observed in the I-SPY2 clinical trial, with p-values less than 0.05 (one-sided Wilcoxon signed-rank test). Notably, based on the predictions from the XGBoost models, TDM1/P, Pembrolizumab (Pembro), and Vincristine (VC) demonstrated pCR rates of 0.784, 0.740, and 0.642, respectively, all significantly surpassing the clinical pCR rates of I-SPY2, which were 0.577, 0.449, and 0.380, respectively. The improvements in pCR rates for the nine arms, excluding Pertuzumab, as predicted by the XGBoost models, range from 0.099 to 0.291.

**Table 3.**
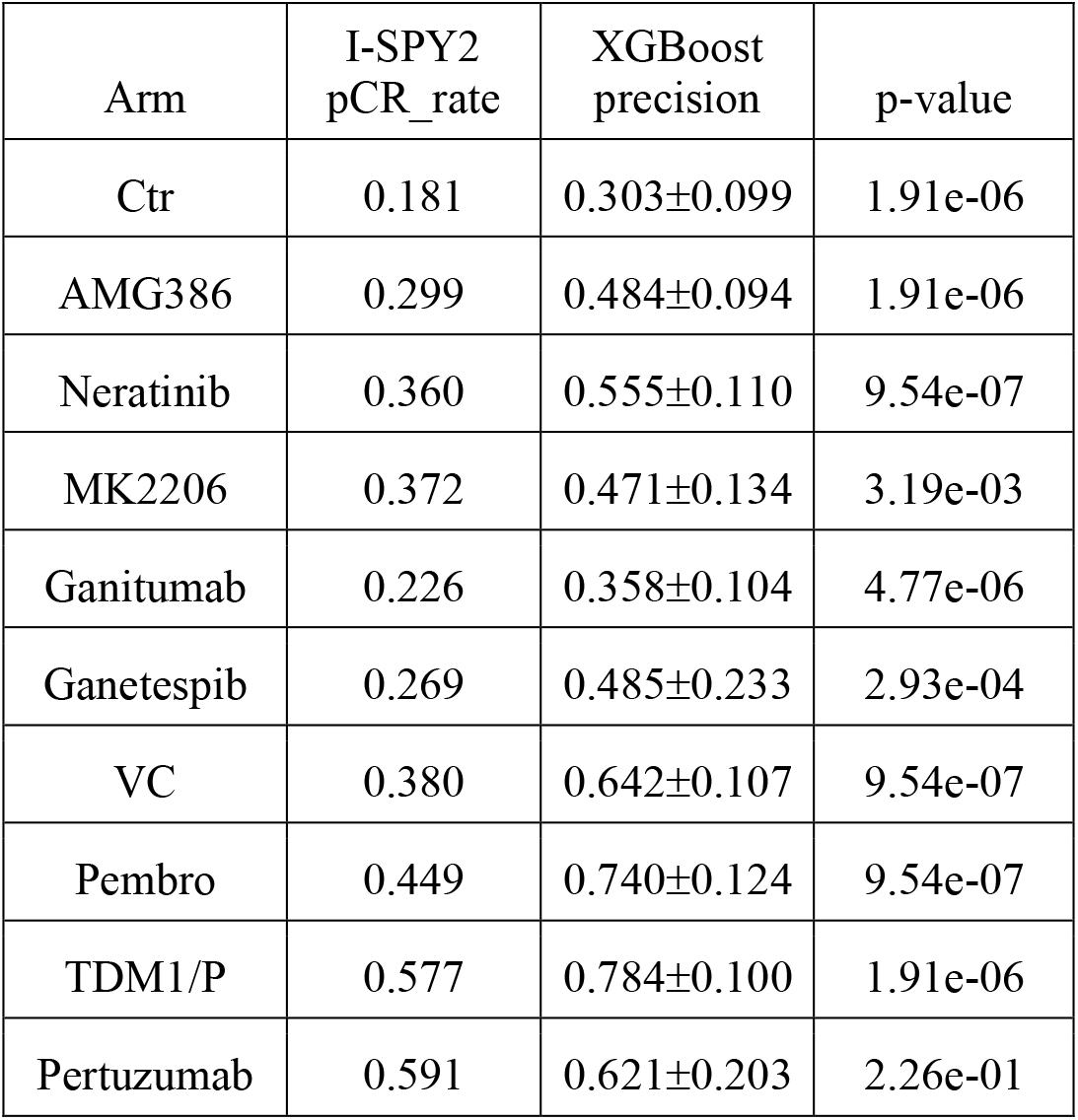
Comparison of pCR rates predicted by XGBoost models and those of I-SPY2.

### ML Predictions Maximize pCR

We applied XGBoost models for the eight arms,excluding Ctr and Neratinib, to the data of the two arms (n=324), Ctr and Neratinib, to predict pCR of each patient to the drugs in the eight arms. Figure 2 presents histograms of the pCR probabilities for each arm. It is important to note that the XGBoost models for Ganitumab, Ganetespib, Vincristine (VC), and Pembrolizumab (Pembro) were trained using data from HER2-patients, and thus were applied to the 228 HER2-samples within the 324 total samples. Similarly, the models for TDM1/P and Pertuzumab, which were trained on HER2+ patient data, were applied to 96 HER2+ samples out of the 324. As illustrated in Figure 2, a substantial number of patients exhibit high pCR probabilities for Pembro, TDM1/P, Pertuzumab, and VC, while the remaining arms show lower probabilities. This trend aligns with the findings in Table 3, where the pCR rates for the first four arms are significantly higher than those for the latter four.

**Figure 2.**
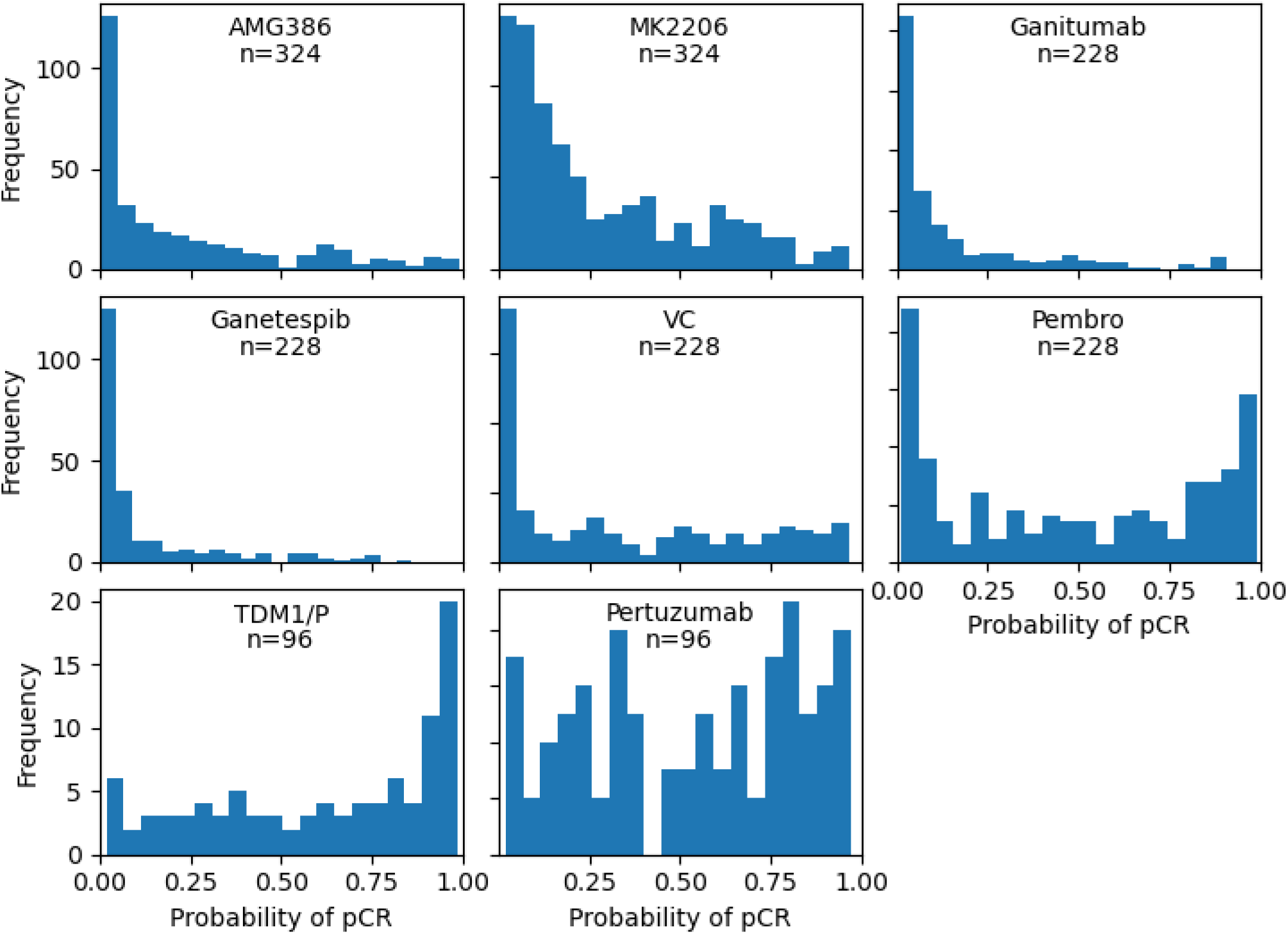
Histograms of the pCR probabilities predicted by XGBoost models for 8 I-SPY2 arms.

To predict whether a patient achieves pCR based on the probabilities shown in Figure 2, we first need to establish a probability threshold. A higher probability threshold will result in fewer patients being predicted to achieve pCR, but those predicted will have a higher pCR rate. To identify the appropriate threshold, we utilized the PPT curve from an XGBoost model specifically trained and tested for one of the I-SPY2 arms. Figure 3 illustrates the PPT curves for the eight I-SPY2 arms. For models with strong predictive power, such as those for Pembro and TDM1/P, a relatively low probability threshold can yield a high pCR rate. Conversely, for models with weaker predictive power, such as those for Neratinib, MK2206, Ganitumab, and Ganetespib, a much higher threshold is required to achieve a satisfactory pCR rate.

**Figure 3.**
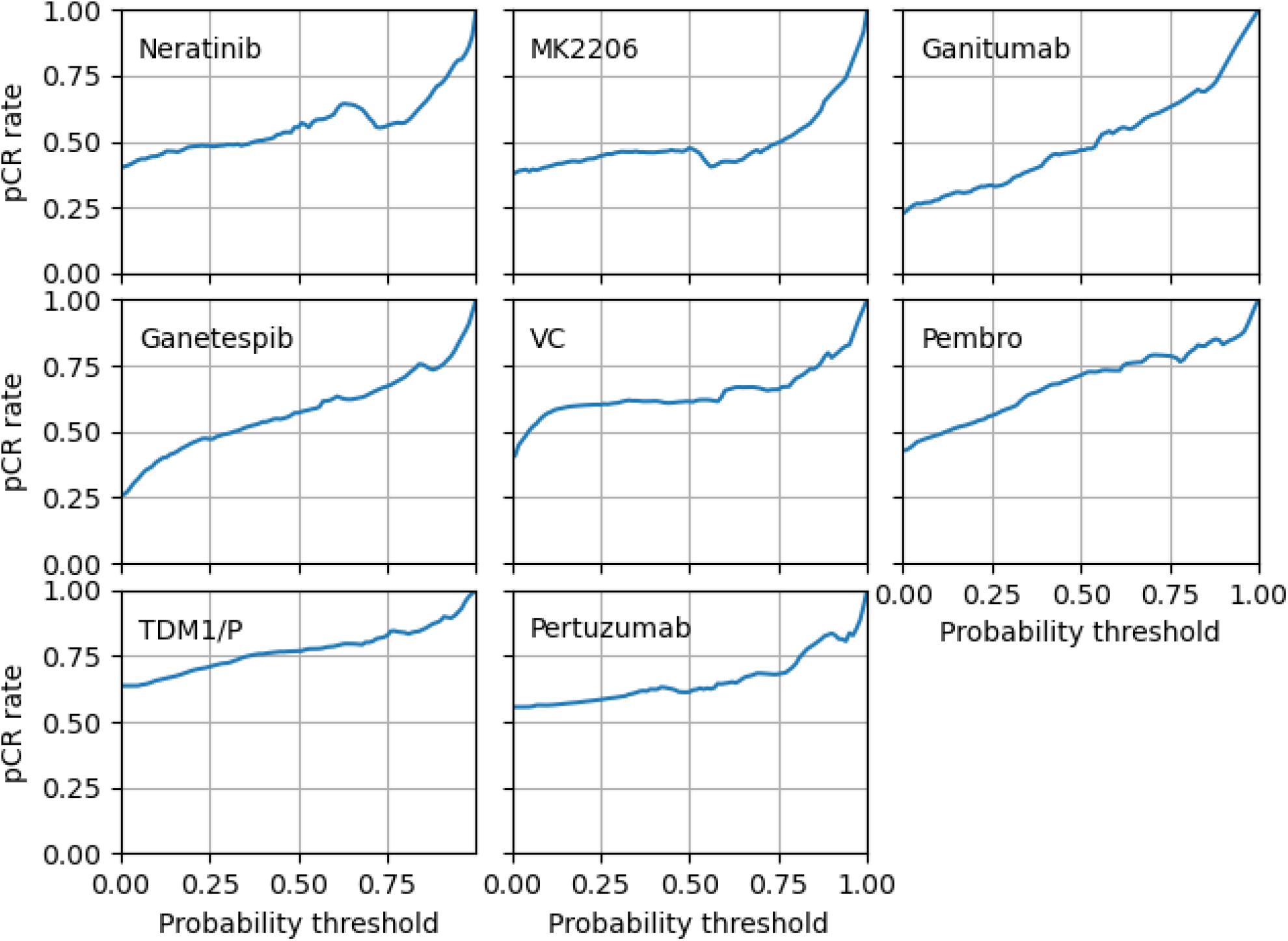
The pCR rate versus the probability threshold of the XGBoost classifiers for the eight I-SPY2

Therefore, based on a target pCR rate, say 0.7, we determined the probability threshold for each arm using its respective PPT curve. We then applied these thresholds to the predicted pCR probabilities from Figure 2 to assess whether a patient would be likely to achieve pCR.

**Table 4** presents the number of patients predicted to achieve pCR for each arm at several target pCR rates. Taking the union across all arms, we also calculated the total number of patients anticipated to achieve pCR in at least one arm. At a target pCR rate of 0.8, 125 out of 324 patients were predicted to achieve pCR, with an actual count of 100 patients who would attain pCR if treated. Similarly, at a target pCR rate of 0.5, 268 patients were predicted to achieve pCR, of which 134 would realistically be expected to do so. Among the four target pCR rates listed in Table 4, a rate of 0.6 seems to be optimal, as it results in the highest expected number of patients (147 of 245 patients) to achieve pCR.

**Table 4.** The number of patients predicted to achieve pCR at different target pCR rates.

| pCR rate | 0.5 | 0.6 | 0.7 | 0.8 |
| --- | --- | --- | --- | --- |
| AMG386 | 20 | 11 | 2 | 1 |
| MK2206 | 19 | 10 | 5 | 2 |
| Ganitumab | 16 | 9 | 5 | 0 |
| Ganetespib | 30 | 12 | 1 | 0 |
| VC | 155 | 125 | 31 | 11 |
| Pembro | 165 | 136 | 113 | 69 |
| TDM1/P | 96 | 96 | 80 | 50 |
| Pertuzumab | 96 | 67 | 26 | 14 |
| All | 268 | 245 | 199 | 125 |

### Genes Associated with pCR

We identified genes that are associated with pCR for each of ten I-SPY2 arms based on the importance score of each feature output from the XGBoost model. The top 100 genes for each arm are listed in the first spread sheet of Supplementary file 1. Since many genes appear in multiple arms, there are 220 unique genes across ten arms, among which 48 genes appear in all ten arms as listed in the second spread sheet of Supplementary file 2. This is not surprising, because nine of the ten arms except TDM1/P include paclitaxel. Figure 4 depicts the expression values of top ten genes in each arm. While nine of the ten genes in the Ctr arm are differentially expressed in pCR and non-pCR groups, only one gene in the MK2206 arm is differentially expressed. The expression levels of some genes were associated with pCR in multiple arms, including lower expression of ABDH1 (ctr, amg386, pembro, pertuzumab), AMZ1 (AMG386, VC, pembro), BAIAP3 (ctr, amg386, Ganetespib, pembro, TDM1/P) and SYTL4 (Ctr, AMG386, Neratinib, VC, Pembro) and higher expression of DENND1C (AMG386, neratinib, pembro), HMGB3 (Ganitumab, VC, TDM1/P, Pertuzumab), HMMR (Ganitumab, TDM1/P, Pertuzumab), PLEKHF1(Ctr, AMG386, Ganitumab,VC, Pembro) and RASEF (AMG386, Ganitumab, Ganetespib, VC, Pembro) (Supplemental file 1).

**Figure 4.**
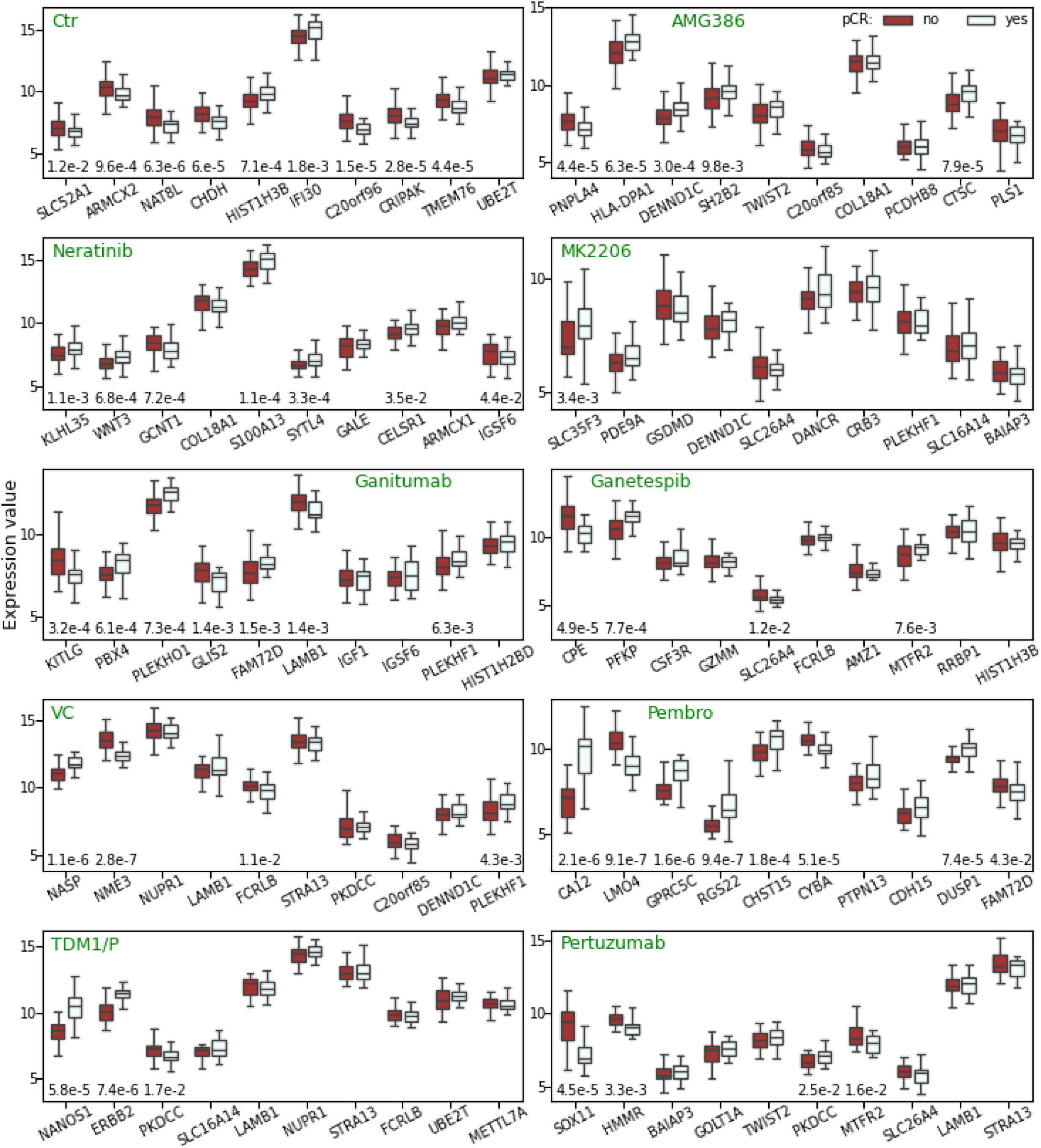
Expression values of top 10 genes associated with pCR in each I-SPY2 arm. The p-values for the comparison between the response and non-response groups that are less than 0.05 are displayed at the bottom of each figure, while those greater than 0.05 are omitted. The Wilcoxon rank-sum test was employed to calculate these p-values.

Of note, since the XGBoost model can capture the nonlinear effect of an input feature on the output target, it can identify genes influencing pCR that are not differentially expressed. GO enrichment analysis identified several GO terms enriched in the top 100 genes. These GO terms are presented in the third spread sheet of Supplementary file 2. The following GO terms appear in multiple arms: endothelial cell morphogenesis (GO:0001886), positive regulation of vascular smooth muscle cell proliferation (GO:1904707), regulation of neuroinflammatory response (GO:0150077), exocytosis (GO:0006887), and nucleosome (GO:0000786). The top five genes in the Pembro arm and the TDM1/P arm are all related to tumorigenesis based on the evidences in the literature (Supplemental file 2).

## Discussion

Human BC is generally categorized into four subtypes based on HR and HER2 status: HR+/HER2, HR-/HER2-, HR+/HER2+, HR-/HER2+ (22, 23), where the HR status is determined by the expression of ER, PR, or both. The HR/HER2 subtyping effectively predicts responses to endocrine and HER2-targeted agents (24). However, with the recent emergence of new targeted therapies for BC, which may function through mechanisms beyond those captured by HR and HER2 expression, a novel RPS-5 subtyping has been proposed (12), which provides better prediction of pCR and may facilitate more effective agent selection for BC treatment. The RPS-5 subtyping relies on five biomarkers: HER2, Immune, DRD, BP-HER2_or_Basal, and BP-Luminal. The Immune and DRD statuses are derived from scores of multiple gene signatures, respectively, while BP-HER2_or_Basal and BP-Luminal classifications are based on BluePrint molecular subtypes (12).

Instead of relying on biomarkers that condense the expression data of multiple genes into a single score, we utilized the expression values of 4,000 genes with the highest variance to train an ML model for predicting pCR. This approach allows us to leverage the comprehensive information within the transcriptome, potentially enhancing prediction accuracy. Our ML models significantly improved the pCR rates for nine out of the ten I-SPY2 arms, except for Pertuzumab. They achieved the highest pCR rates for the Pembro and TDM1/P arms at 0.74 and 0.78, with sensitivities of 0.73 and 0.71, respectively. We also explored two additional ML models, support vector machine and logistic regression with elastic-net regularization, but their predictive performance was inferior to XGBoost. Consequently, their results are not included.

We have trained ten XGBoost models to predict pCR for ten drug combinations. If gene expression and pCR data for additional treatments become available, we can similarly train XGBoost models for those therapies. The challenge is effectively utilizing these predictions to select the best drug for individual patients, as the models show varying predictive power shown in Tables 2 and 3. A fixed probability threshold of 0.5 for pCR classification may not be optimal, especially if multiple drugs predict pCR. To address this, we developed a drug selection strategy based on the PPT curves of the XGBoost classifiers. We identify a probability threshold from the PPT curve that aligns with the desired pCR rate. For patients predicted to achieve pCR with multiple drugs, we can choose the drug with fewer side effects or the highest sensitivity at the threshold. This strategy ensures a target pCR while selecting the most appropriate drug for each patient.

While predicting pCR is crucial for determining treatment strategies, identifying genes linked to pCR, is equally important to finding genes as potential targets for intervention for drug development. Our XGBoost models leverage importance scores to assess the influence of individual genes on pCR, pinpointing key genes that significantly impact this outcome. Analysis of the top five genes identified for Prembo and TDM1/P reveals that all involved in tumor development and progression, underscoring the relevance of the genes identified through our XGBoost models.

A major limitation of this study is the small sample size. While our XGBoost models demonstrated significantly higher precision, which is equivalent to the pCR rate, compared to the I-SPY2 trial across nine arms excluding Pertuzumab, the standard error in predicted pCR rates remains high due to the limited sample. A larger sample could reduce the standard error, enhancing model robustness and potentially improving predictive power as indicated by metrics like AUC and F1 score. Despite this, the study underscores the potential of ML models trained on gene expression data to provide accurate pCR predictions for BC patients.

## Data Availability

All data produced in the present work are contained in the manuscript.

https://github.com/CaixdLab/BRCA_NATpCR

## Supplemental information

Supplemental file1 (xlsx) Supplemental file 2 (word)

## Acknowledgement

We thank Dr. Xiaodong Cai at the College of Engineering of University of Miami for assistance on methodology and data analysis.

## Author Contributions

ZC conceived the study. Both authors contributed to methodology and data analysis. JC wrote the computer programs for implementing the machine learning methods and data visualization. Both authors wrote, reviewed, and approved the manuscript.

## Data availability

This study used the gene expression and clinical data from the I-SPY2 clinical trial (12). The microarray gene expression data can be accessed in GEO under the accession number GSE194040. The clinical data are provided in Table S2 of (12) available on the journal’s website. The computer programs developed for this study are available on GitHub (https://github.com/CaixdLab/BRCA_NATpCR), allowing for the replication of all results presented in this paper.

