## Supplemental file 2 for "Machine Learning for Predicting and Maximizing the Response of Breast Cancer Patients to Neoadjuvant Therapy"

**Supplementary information**

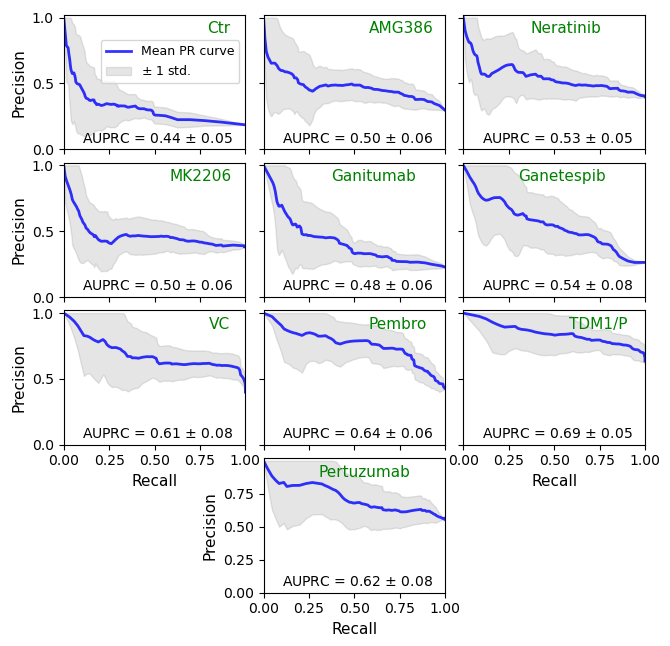

Figure S1 Precision-recall curve of XGBoost classifiers for predicting the pCR in ten arms of I-SPY2.

**Role of top genes in tumorigenesis**

Supplementary file 2 lists top 100 genes that are associated with pCR for each of 10 I-SPY2 arms. To see if these genes are relevant in breast cancer, we completed a literature search about the information of top five genes of two arms that have highest predive power for pCR, Pembro and TDM1/P. The top five genes of Pembro are CA12, LMO4, GPRC5C, RGS22, and CHST15. CA12 is robustly regulated by estrogen via ER alpha in BC cells (1). High expression of CA12 is linked to better overall survival, a lower risk of metastasis, and less aggressive tumor features in invasive breast carcinoma (2, 3). LMO4 is overexpressed in a significant percentage of human BC and cell lines (4), and LMO4 overexpression may disrupt the normal tumor suppressor activities of Tip, promoting BC progression (5). GPRC5C is one of the genes identified as being regulated by estrogen in BC cells (6). RGS22 was found to be expressed in several cancers of epithelial origin, and overexpression of RGS22 reduced cell migration and invasive potential in a highly metastatic esophageal cancer cell line (7). CHST15 expression directly contributes to the invasive and metastatic potential of BC cells (8). Therefore, all five top genes of the Prembro arm are related to cancer development.

The top five genes of TDM1/P are NANOS1, HER2, PKDCC, SLC16A14, and LAMB1. The NANOS1 gene is associated with BC progression and invasiveness, playing a pivotal role in epithelial-mesenchymal transition (EMT), since its expression is inversely correlated with E-cadherin (CDH1) expression (9, 10). HER2 amplification is the primary pathway of HER2 receptor overexpression and is a major driver of tumor development and progression in a subset of BCs (11). The gene PKDCC (also known as VLK) is a secreted kinase that has been shown to promote tumor processes like cell invasion and colony formation by phosphorylating cell surface proteins (12). SLC16A14 is a member of the SLC16 gene family, which is associated with monocarboxylate transporters. Pan-cancer analysis has suggested that other members of the SLC16 family, like SLC16A2 and SLC16A9, play a role in BC and may be linked to poor prognosis (13). The LAMB1 gene was found to promote proliferation and metastasis of gastric cancer and nasopharyngeal carcinoma (14, 15). Based on these findings, all top five genes of the TDM1/P arm are related to tumorigenesis.
